# Treatment effect variation in brain stimulation across psychiatric disorders

**DOI:** 10.1101/2020.05.02.20088831

**Authors:** Stephanie Winkelbeiner, Whitney Muscat, Andrea Joanlanne, Nikolaos Marousis, Stefan Vetter, Erich Seifritz, Thomas Dierks, Philipp Homan

## Abstract

Noninvasive brain stimulation methods such as transcranial magnetic stimulation (TMS) and transcranial direct current stimulation (tDCS) are promising add-on treatments for a number of psychiatric conditions. Yet, some of the initial excitement is wearing off. Randomized controlled trials (RCT) have found inconsistent results. This inconsistency is suspected to be the consequence of variation in treatment effects and solvable by identifying responders in RCTs and individualizing treatment. However, is there enough evidence from RCTs that patients do indeed respond differently to treatment? This question can be addressed by comparing the variability in the active stimulation group with the variability in the sham group across studies.

We searched MEDLINE/PubMed and included all double-blinded, sham-controlled RCTs and crossover trials that used TMS or tDCS in adults with a unipolar or bipolar depression, bipolar disorder, schizophrenia spectrum disorder, or obsessive compulsive disorder. In accordance with the PRISMA guidelines to ensure data quality and validity, we extracted a measure of variability of the primary outcome.

A total of 114 studies with 5005 patients were considered in the analysis. We calculated variance-weighted variability ratios for each comparison of active versus sham stimulation and entered them into a random-effects model. We hypothesized that treatment effect variation in TMS or tDCS would be reflected by increased variability after active compared with sham stimulation, or in other words, a variability ratio greater than one.

Across diagnoses, we found a slight increase in variability after active stimulation compared with sham (variability ratio = 1.05; 95% CI, 1.01-1.11, P = 0.012). This effect was likely driven by studies in patients with schizophrenia who received rTMS compared with sham (variability ratio = 1.11; 95% CI, 1.03-1.2, P = 0.007).

In conclusion, this study found evidence for treatment effect variation in brain stimulation, particularly for studies in schizophrenia. The extent of this variation, however, was modest, suggesting that the need for personalized or stratified medicine is still an open question.

## Introduction

The emergence of noninvasive brain stimulation for the treatment of various psychiatric conditions has brought promising possibilities. Stimulation methods such as transcranial magnetic stimulation (TMS) and transcranial direct current stimulation (tDCS) have been used as addon treatment^1^ and have convinced with their minimal risk profile,^2^ the prospect of noninvasively interfering with neuronal transmission, and the easy-to-use application. Popularity, particularly for TMS, increased considerably after the Food and Drug Administration’s^3^ approval of its application as an add-on to the conventional therapy for major depressive disorder in 2008 and obsessive-compulsive disorder (OCD) in 2018.

Consistent evidence for the superiority of TMS or tDCS over sham for other psychiatric disorders is lacking so far. Positive symptoms in schizophrenia are one such example: the initially promising results^4–6^ developed into a more heterogeneous picture over the years.^7^ One explanation for this might be the drop in effect sizes over time.^8,9^ Another reason, often brought up by both researchers and clinicians (including ourselves^10^), is to assume variation in treatment effects in brain stimulation.^11^ This assumption has motivated the idea to personalize medicine by identifying responders to treatment through predictive biomarkers.^12–15^

However, the question remains whether there is enough evidence to conclude that patients do indeed differ in their response to noninvasive brain stimulation. In addition, the extent of such variation would be important as well, as it would determine the corresponding need for personalized psychiatry. One way to address this question is by comparing the variance between treatment and control groups of randomized, controlled trials (RCT).^16^ Greater variability in the active stimulation group would indicate that there is a component of variation, the patient-by-treatment or subgroup-by-treatment interaction, indicating variability of treatment effects.^17^ This approach has recently been used in the context of antipsychotics^18^ and antidepressants.^19–22^ Perhaps surprisingly, most of these meta-analyses found little evidence for treatment effect variation.^18–20,22^

To evaluate the heterogeneity in treatment effects for noninvasive brain stimulation (TMS and tDCS), we used the same approach of examining variability ratios in RCTs with patients across psychiatric diagnoses (unipolar depression, schizophrenia specrtum disoder, OCD, and bipolar disorder). We hypothesized that (1) the often claimed treatment effect variation in brain stimulation would be indicated by increased variability after active stimulation compared with sham, reflected by an overall variability ratio (VR) of greater than one. Further, we hypothesized that (2) the variability ratio is not dependent on the diagnostic group or (3) stimulation method.

## Methods

### Selection criteria

We included all studies that met the following eligibility criteria: (1) study population of adult patients with either a diagnosis of unipolar or bipolar depression, schizophrenia spectrum disorder, OCD, or bipolar disorder; (2) using noninvasive brain stimulation including TMS and tDCS; (3) RCTs or crossover trials; (4) published in a peer-reviewed English journal; (5) no case reports, case series, opinion pieces; (6) no animal research.

### Search strategy

We conducted a comprehensive search of the electronic database MEDLINE (PubMed.gov) from January 1999 - October 2018 with the following combination of keywords: “depression” or “MDD” or “unipolar” and “transcranial magnetic stimulation” or “rTMS” or “TMS” or “transcranial direct current stimulation” or “tDCS”; “bipolar” or “mania” and “transcranial magnetic stimulation” or “rTMS” or “TMS” or “transcranial direct current stimulation” or “tDCS”; “obsessive compulsive disorder” or “OCD” or “obsession” or “compulsion” and “transcranial magnetic stimulation” or “rTMS” or “TMS” or “transcranial direct current stimulation” or “tDCS”; “psychosis” or “schizophrenia” or “positive symptoms” or “auditory hallucinations” or “thought disorder” or “delusions” or “hallucinations” or “thinking” or “disorganization” and “transcranial magnetic stimulation” or “rTMS” or “TMS” or “transcranial direct current stimulation” or “tDCS”. The only search filters that were applied were “human”, and “English language”. In addition to the electronic database search, we searched the references of recent reviews and meta-analyses.^23–27^

### Data extraction

In adherence with the PRISMA guidelines,^28^ two independent researchers (A.J. and W.M.) conducted the literature search and screened the articles, eliminated duplicates, decided whether the article met the inclusion criteria, and extracted all relevant data from the final articles (Supplementary Figure 1). Their independent searches were compared by S.W., discrepancies investigated, and resolved by discussion. The risk of bias of the included studies was assessed independently by S.W. and N.M. according to the Cochrane Handbook for Systematic Reviews of Interventions.^29^

We extracted an available variance measure (standard deviation, standard error, or confidence interval) and means for the primary outcome measure at baseline and outcome for the active and sham group. Additionally, we extracted information on the study design, sample size, participants’ characteristics (diagnosis, treated symptom, sex, and age), stimulation parameters (targeted hemisphere, number of sessions, and whether neuronavigation was used), and the effect size. For TMS studies, we extracted also target location and stimulation frequency. For tDCS studies, we extracted also electrode placement of the anode and cathode and stimulation intensity.

Note that in cases of multi-arm RCTs, we omited study arms that were not relevant to the question of this meta-analysis.^30^ For example, we omitted the arm of one study^31^ in which TMS as an add-on to sertralin was compared to sham, while we included the arm in which TMS as a standalone treatment was compared to sham.

### Data preprocessing

The majority of the included studies were RCTs (87%), with crossover trials making up only a small percentage (13%). Nevertheless, the inclusion of crossover trials required additional considerations.^30^ We decided to exclude the second period of each crossover trial thus treating the trial as if it was a parallel trial. To check that this approach did not lead to biased results we conducted sensitivity analyses including also the second period of crossover trials (Supplementary Figure 2). Of note, variance measures and means had to be estimated from figures for 17 (12%) studies. In the case of multi-arm trials, in which more than one arm of the RCT was considered relevant to this meta-analysis,^31–49^ we divided the sample size of the sham group by the number of intervention arms while retaining standard deviation (*s*) and mean.^30^ This allowed us to create multiple pair-wise comparisons for those studies.

For the analysis, we used the pre-post difference scores to account for baseline differences. For the 121 (88 %) studies that reported raw outcome scores, we calculated the mean pre-post difference score (M_Δ_) with

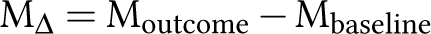

The calculation of the respective standard deviation (*s*_Δ_) is less straightforward with^50,51^

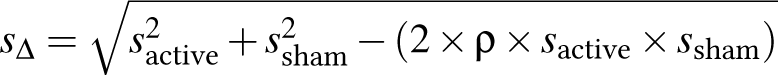

and required an approximation for the overall correlation coefficient (*ρ*). Thus, we used the 17 (12 %) studies that had reported the standard deviation for the active and sham groups for baseline (*t*_0_), outcome (*t*_1_), and pre-post difference scores to calculate the correlation coefficient for the active (ρ_active_) and sham (ρ_sham_) group, respectively, with

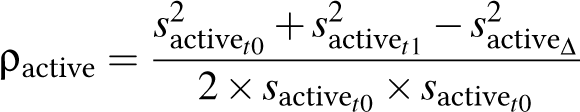

Next, we averaged over ρ_active_ and ρ_sham_ to obtain one correlation coefficient per comparison. Finally, we averaged across the 17 comparisons and used this average coefficient (ρ_average_ = 0.57) for imputation in the calculation of *s*_Δ_.^29,30^ While this approach was possible for the majority of studies, there were 3 studies that had reported the standard deviation of the percentage pre-post difference scores^52–54^ and one that had reported the range of the pre-post difference scores^55^ but provided no information on the raw outcome scores. Therefore, those studies had to be excluded.

### Statistical analysis

The variance of the active groups comprises the same variance components (within-patient variation, regression to the mean, and measurement error) as the sham group, with the only difference that the active group can comprise also a treatment-by-patient or treatment-by-subgroup interaction. This variance component reflects response differences between patients or subgroups. Thus, in the case of individual response difference, we would expect an increased variation (VR > 1) in the active group compared with sham. To test this, we calculated the VR for each comparison of active stimulation (TMS or tDCS) versus sham as log variability ratio (lnVR)^56^ with

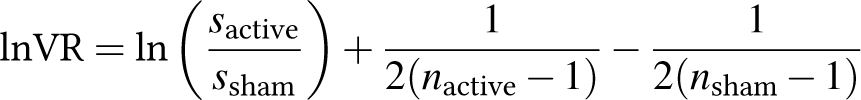

where *s*_active_ and *s*_sham_ were the standard deviations of the pre-post difference scores, and *n*_active_ and *n*_sham_ the respective sample sizes.^16^ The corresponding sampling variance 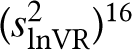 for each study was defined as

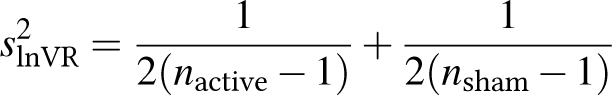

We weighted each lnVR with the inverse of its 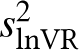.^57^ To quantify the true individual response, after adjusting for within-patient variability and regression to the mean,^17,58^ we fitted a random-effects model stratified by diagnostic group. For better interpretability, we transformed the results back from the log scale: VR greater than 1 indicating greater variability under active stimulation compared with sham and a VR smaller than 1 indicating less variability under active stimulation compared with sham.

Finally, we calculated the standardized mean difference (SMD) as a general effect size measure to compare our analysis with previous reports. By taking the log of the ratio of means, the SMD was centered symmetrically around zero, yielding Hedges’ *g*.^59^ We also plotted the SMDs against year of publication stratified by diagnostic group to examine the relationship of the effect sizes and time.

### Data and code availability

All analyses were performed in R (version 3.5.2),^60^ the calculation of VR and SMD was done with the R package *metafor* (version 2.4.0).^59^ This paper was written using *knitr* (version 1.28)^61^ in RStudio (version 1.2.50 42).^62^ All data and code are freely available online to ensure reproducibility (https://osf.io/6w947/). This study was pre-registered on the Open Science Forum platform (https://osf.io/8uxec).

## Results

### Descriptive statistics

A total of 114 were included that investigated treatment with active TMS or tDCS in depression,^31,31,33,35,37–43,45–49,63–108^ schizophrenia,^5,6,32,34,44,109–134^ OCD,^36,135–145^ and bipolar disorder^40,146–151^ compared with sham over the last 22 years. Some studies compared more than one active stimulation to sham which is why we considered 138 comparisons in this meta-analysis. We had to exclude studies that failed to report the necessary data to calculate pre-post difference scores (4 studies^52–55^) and that defined more than one primary outcome measure (1 study^152^). We excluded the second period of 12 crossover trials^65,66,72,76,80,100,118,121,136,138,145,153^) and dropped 1 study’s arm^31^ that was irrelevant for this meta-analysis. Importantly, none of the included studies used a design such as a repeated crossover trial that would have allowed to estimate individual response directly.^154^

Overall, a total of 5005 patients were included. Of these 3240 (65%) had a diagnosis of unipolar or bipolar depression, 1262 (25%) schizophrenia spectrum disorder, 278 (6%) OCD, and 224 (4%) bipolar disorder. Further, 99 (87%) comparisons investigated TMS and 15 (13%) tDCS compared with sham. Our evaluation of the risk of bias showed that 61 (44%) comparisons had a high risk of bias, while only 22 (16%) a low risk. For 55 (40%) comparisons the risk was unclear (see Supplementary Figure 3). Therefore, we conducted sensitivity analyses to investigate the reliability of our results (see Supplementary Figure 4). For more details on the included studies see Supplementary Figure 5 and the Supplementary Tables 1-14.

### Variability ratio

We found an overall slightly increased variability after active stimulation compared with sham across diagnostic groups and stimulation methods (VR = 1.05, 95% CI: 1.01, 1.1, P = 0.014, Figure 1). This effect was most likely driven by studies in patients with schizophrenia (VR = 1.1, 95% CI: 1.02, 1.19, P = 0.014, Figure 1). The subgroup analyses by stimulation method showed that only for TMS studies in schizophrenia there was a 1% increase in variability evident after active compared with sham stimulation (Supplementary Figures 6-9). For all other diagnoses, there was no evidence for increased variability, irrespective of diagnostic group and stimulation method. We estimated that 1% of the total variance was due to heterogeneity in true effects and not attributable to sampling variance.

**Figure 1.**
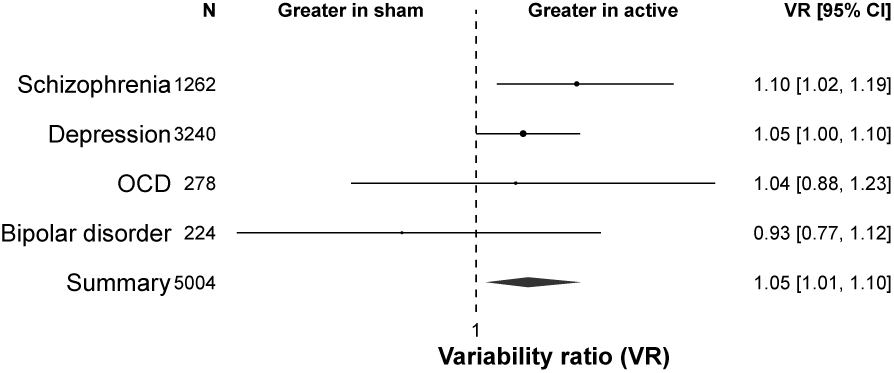
Variability ratio by disorder. The forest plot shows the VR together with its 95% confidence interval (CI) for active stimulation vs sham by disorder. Within the diagnostic groups, the variance was not significantly increased by the stimulation (TMS or tDCS) compared with sham, except for studies in patients on the schizophrenia spectrum.

### Mean effect size

Additionally, we calculated the standardized mean differences to obtain an index of the effectiveness of brain stimulation. Overall, we found a medium to large effect size (SMD = 0.62, 95% CI: 0.32, 0.92, P < 0.001; Figure 2) across diagnostic groups. This indicates that noninvasive brain stimulation was on average more effective than sham and is in line with previous meta-analyses in depression (TMS: odds ratios between 1.69 and 7.44^73,155^; tDCS: odds ratio = 4.17^155^), schizophrenia (Hedges’ g between 0.39 and 0.63^156,157^), and OCD (Hedges’ *g* = 2.94^158^).

**Figure 2.**
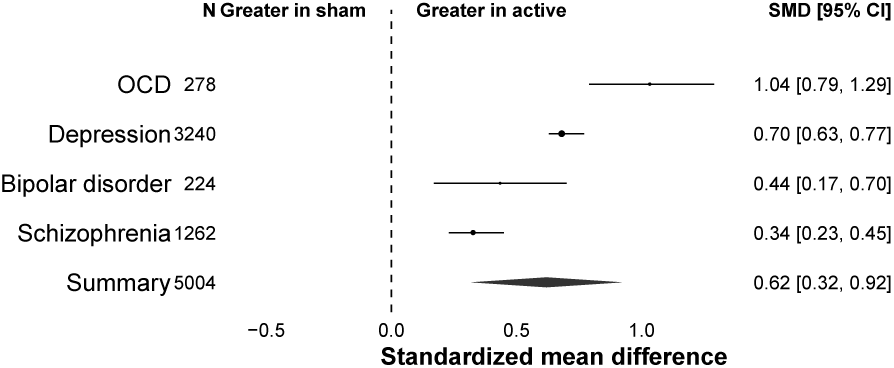
Standardized mean difference. The forest plot shows the standardized mean difference (SMD) together with its 95% confidence interval (CI) for active stimulation vs sham by disorder. The overall effect size was greater for active stimulation compared with sham for all disorders.

Further, we found an inverse relationship of effect sizes and year of publication for the studies in patients with a schizophrenia spectrum diagnosis, but not for any of the other diagnostic groups (Supplementary Figure 10).

## Discussion

The observed variation in clinical trials and clinical practice is often viewed as proof that patients differ in their response to noninvasive brain stimulation. If this was true, one would expect that active stimulation should increase variability compared with sham stimulation. The empirical evidence in this study does indeed provide some support for this assumption, although our study could not determine whether this was due to differences between individuals or between subgroups, as both would lead to increased overall variability compared with sham. Yet, the size of this variability increase was modest and mostly driven by treatment effect variation in schizophrenia trials and to a smaller extent in depression trials. Consistent with previous meta-analyses, our study also found support for the overall efficacy of brain stimulation across disorders, even though a substantial risk of bias because of the mostly small studies has to be acknowledged.

Our findings lend at least some support for the recent trend in psychiatry to personalize treatment^159,160^ in general and brain stimulation in particular.^12,161^ One reason for this trend might be that the initally promising potential of noninvasive brain stimulation^162^ for the treatment of psychiatric and neurological conditions,^24,25,27,73,155,157,158^ has started to wear off. Despite medium to large effect sizes from depression trials,^99,163^ the promised beneficial effects did not appear to show in all patients. While some improved, others appeared to remain unchanged or even deteriorated. These oberved outcome differences leave patients and clinicans unsatisfied, and researchers tempted to assume variation in treatment effects. Naturally, this has motivated the search for biomarkers to identify those patients who are likely to respond more favorably to treatment and tailor treatments accordingly. Yet, despite encouraging attempts,^12–14^ (that occasionally did receive some pushback^164,165^) so far no reliable and clinically applicable biomarkers have been identified for treatment response prediction in individuals or subgroups.

One of the challenges in predicting treatment response might be rooted in the assumption that treatment effects vary in clinical trials and clinical practice. More often than not is this assumption the result of a somewhat arbitrary distinction between “responders” and “nonresponders”.^166,167^ Yet, the presence of other variance components that might explain a majority of the outcome variance is sometimes overlooked.^168^ Thus, evaluating variances can give a first hint about whether a personal or subgroup element of response is indeed present in the data.^16^ Our results provide some evidence for the presence of such treatment effect variation, driven mostly by studies in patients on the schizophrenia spectrum who were treated with TMS. For those studies, variability was slightly increased after TMS compared with sham. This deviates from similar recent meta-analyses of antipsychotic drug trials^18,169^ and antidepressant drug trials,^19,20,22^ where little evidence for treatment effect variation was found.

Notably, our results for the overall mean effect size of brain stimulation is largely in line with previous meta-analyses in depression (TMS: odds ratios between 1.69 and 7.44 ^73,155^; tDCS: odds ratio = 4.17^155^), schizophrenia (Hedges’ *g* between 0.39 and 0.63^156,157^), and OCD (Hedges’ *g* = 2.94^158^). In addition, we found a negative correlation of effect size and publication year for schizophrenia studies, which was also in line with previous studies.^8,9^ This effect might be explained by the larger sample sizes (N > 70) of more recent studies,^128,134^ while the initial RCTs were performed in treatment groups of 10 or less patients (16 comparisons,^41,42,45,65,66,80,82,131,132,138,141,142,145^ 12%). Such small sample studies have low average statistical power,^170,171^ which can be consequential. Chances are lower to detect the “true” effect that actually reflects a statistically significant result. Thus, effect sizes are overestimated which reduces chances of reproducibility.

What are the clinical implications of our results for the treatment with noninvasive brain stimulation? This metaanalysis of variance differences from RCTs has provided some evidence to expect individual or subgroup differences in response to brain stimulation. One explanation might be the presence of subgroups that might have responded differently to brain stimulation with the more severely ill patients improving less or not at all and the less ill patients improving more.^17^ Such a case would argue for “stratified medicine” rather than “personalized medicine”, in which subgroups of patients receive varying treatments. Thus, the analysis of variability ratios does not allow to distinguish between those cases, i.e. patient-by-treatment and subgroup-by-treatment interactions.^172^ However, although we did find evidence for treatment effect variation, the overall extent of such variation was modest and most pronounced for schizophrenia, and even for schizophrenia the confidence intervall ranged from 2% to 19%, indicating that the need for personalized or stratified medicine, respectively, could be anything from minimal to moderate. It is therefore an open question whether these findings are strong enough to warrant the search for predictive biomarkers, no matter how promising the approach.^161,173,174^

## Limitations

Some limitations merit comment. First, we decided to calculate the variability ratio despite a significant mean-variance relationship for studies in patients on the schizophrenia spectrum (Supplementary Figure 11). This may limit the variability ratio in its applicability to identify shifts in variances and would call for the use of the coefficient of variation ratio (CVR) instead.^16^ Yet, the CVR is limited itself by the fact that the logarithm can only be taken from positive values. Thus, negative mean values cannot be dealt with without losing information. Using pre-post difference scores can result in negative values in the case of worsening of symptoms 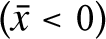 or zero values in the case of no symptom change 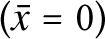 from baseline to outcome. More importantly, the CVR is scale dependent. For values on the interval scales with a mean close to zero but a standard deviation far from zero, the CVR becomes arbitrarily large and the mean-variance proportionality which makes the CVR meaningful is lost. Therefore, the CVR should only be used for outcome data on the ratio scale with a true zero.^175^ Last, the majority of the included studies were rated as having high risk of bias, which suggests a low quality of evidence. This was due to various factors such as not clearly defined primary outcome measures, incomplete or selective data reporting, potentially misleading report of outcome data, and failure to report baseline data. We undertook various measure to account for these shortcomings (see Methods) and excluded the studies that did not report the values needed to infer the necessary values. In addition, we performed sensitivity analyses excluding the studies for which values had been inferred. The results of the sensitivity analyses showed the same direction as the main analyses.

## Conclusion

Although this study did find evidence for treatment effect variation in brain stimulation (particularly for studies in schizophrenia), the extent of this variation was overall modest, suggesting that the need for personalized or stratified medicine is still an open question.

## Data Availability

All data and code are freely available online to ensure reproducibility (https://osf.io/6w947/). This study was pre-registered on the Open Science Forum platform (https://osf.io/8uxec).

## Acknowledgments

The authors thank Majnu John, PhD, Department of Mathematics, Zucker School of Medicine at Northwell/Hofstra, for advice on the analysis of the current study, as well as Anil Malhotra, MD, Department of Psychiatry, Zucker School of Medicine at Northwell/Hofstra, for helpful comments on the manuscript. These individuals received no additional compensation, outside of their usual salary, for their contributions.

